# Diagnostic Yield of Exome Sequencing in a Diverse Pediatric and Prenatal Population is not Associated with Genetic Ancestry

**DOI:** 10.1101/2023.05.19.23290066

**Authors:** Yusuph Mavura, Nuriye Sahin-Hodoglugil, Ugur Hodoglugil, Mark Kvale, Pierre-Marie Martin, Jessica Van Ziffle, W. Patrick Devine, Sara L. Ackerman, Barbara A Koenig, Pui-Yan Kwok, Mary E. Norton, Anne Slavotinek, Neil Risch

## Abstract

**Purpose:** It has been hypothesized that diagnostic yield (DY) from Exome Sequencing (ES) may be lower among patients with non-European ancestries than those with European ancestry. We examined the association of DY with estimated continental genetic ancestry in a racially/ethnically diverse pediatric and prenatal clinical cohort.

**Methods:** Cases (N=845) with suspected genetic disorders underwent ES for diagnosis. Continental genetic ancestry proportions were estimated from the ES data. We compared the distribution of genetic ancestries in positive, negative, and inconclusive cases by Kolmogorov Smirnov tests and linear associations of ancestry with DY by Cochran-Armitage trend tests.

**Results:** We observed no reduction in overall DY associated with any continental genetic ancestry (Africa, America, East Asia, Europe, Middle East, South Asia). However, we observed a relative increase in proportion of autosomal recessive homozygous inheritance versus other inheritance patterns associated with Middle Eastern and South Asian ancestry, due to consanguinity.

**Conclusions:** In this empirical study of ES for undiagnosed pediatric and prenatal genetic conditions, genetic ancestry was not associated with the likelihood of a positive diagnosis, supporting the ethical and equitable use of ES in diagnosis of previously undiagnosed but potentially Mendelian disorders across all ancestral populations.

## Introduction

Advances in Exome Sequencing (ES) technology has led to use of ES in establishing molecular diagnoses for Mendelian diseases in children and adults. This has prompted recommendations for ES as the first line genetic test for certain clinical indications such as neurodevelopmental disorders.^1^ The probability of a positive case classification from ES (diagnostic yield) may differ due to factors such as: number of parents sequenced with proband, parental age, variant of uncertain significance (VUS) calling threshold, consanguinity, clinical indication or phenotype presentation, sex and age of proband, genetic ancestry, or a combination of these factors. Most of the studies on ES diagnostic yield have been conducted in predominantly European ancestry populations.^2^

Relatively little is known about the diagnostic yield (DY) from ES in individuals with ancestry such as African, East Asian, South/Central Asian, Middle Eastern, Native American and Pacific Islander, as well as ancestrally admixed individuals.^3^ Genetic variant data from individuals with non-European ancestry is underrepresented in genetic and genomic databases,^2^ and it has been suggested that DY may be lower in those with non-European ancestry. Some have found higher rates of VUSs in individuals with African, Native American and other underrepresented ancestries compared to those of European ancestry,^4,5^ which suggests the potential for reduced diagnostic yield in non-European ancestry populations.

To investigate this question in the context of ES for rare undiagnosed but suspected Mendelian disorders, we analyzed the association of diagnostic yield with estimated global genetic ancestry in an ancestrally diverse cohort of pediatric and prenatal cases who underwent ES, and how it relates to the self-identified race/ethnicity of the parents of the cases.

Our analysis was based in the Program in Pediatric and Prenatal Genomic Sequencing (P^3^EGS) cohort at the University of California, San Francisco (UCSF), which is part of the Clinical Sequencing Evidence-generating Research (CSER) consortium.^6^ Cases in the P^3^EGS cohort had a wide range of clinical indications for ES, and was ancestrally diverse, with 66% of all parents and 80% of parents providing race/ethnicity information self-identifying as non-white.^7^

The association between diagnostic yield and important factors other than genetic ancestry has been reported previously using the same cohort.^7^

## Materials and Methods

### Study participants, demographics, and inclusion criteria

Pediatric (N=529) and prenatal (N=316) cases and their available biological parents were primarily enrolled at one of five sites in the San Francisco Bay area and Central Valley of California. The five sites included UCSF Benioff Children’s Hospital San Francisco and Benioff Children’s Hospital Oakland, Zuckerberg San Francisco General Hospital, the Betty Irene Moore Women’s Hospital at Mission Bay, and the Community Medical Center in Fresno (See Slavotinek et al., 2023 for full details; inclusion, and exclusion criteria, institutional review board (IRB) approval and consent).

### Self-reported race/ethnicity of parents

Parents of affected probands voluntarily responded to questions about their demographic background on a structured instrument. Details are provided in Supplementary Material.

### Exome sequencing

ES of samples from the probands and available parents was done at the UCSF Genomic Medicine Laboratory (GML). Details of the sequencing, quality control and selection of markers for genetic ancestry analyses are provided in Supplementary Material.

### Genetic ancestry, admixture, relatedness, and consanguinity analysis

Individual genetic ancestry admixture proportions were estimated using the ADMIXTURE software package^8^ using a set of 53,665 exome-wide markers (See Supplementary Material). A supervised approach, whereby unrelated individuals in (K) reference populations are assumed to have 100% reference genetic ancestry, was utilized to estimate the individual genetic admixture proportions in individuals of the P^3^EGS cohort (parents and probands). We created K=7 reference continental ancestral populations from the Human Genome Diversity Panel (HGDP) individuals, based on literature.^9^ The 7 reference populations were: African -Afr (Yoruba, Mandenka N=40), Native American – Amr (Colombia, Karitiana, Surui, Pima, N=40); East Asian -Eas (Han, Japanese N=40); Middle Eastern -Mid (Druze, Palestinian, Bedouin, N=40); European -Eur (French, Orcadian, Tuscan, Sardinian, N=40); Oceanian -Oce (Papuan, Melanesian N= 30); South Asian -Sas (Pathan, Sindhi, N=39).

Principal components analysis (PCA) was also performed on the HGDP samples using the SmartPca program, part of the EIGENSOFT4.2 software package^10^ using the same 53,665 markers. The P^3^EGS samples were then projected onto the HGDP-derived PCs to facilitate geographic interpretation of the P^3^EGS participants.

Genetic kinship between P^3^EGS participants (probands, Parents) was estimated using PC-Relate.^11^ Genetic ancestry was controlled by using the first 8 PCs from PCA in PC-Relate in order to estimate only recent genetic relatedness due to family structure. Linkage disequilibrium (LD) pruning (0.1 kb in a 1000 kb window) was done to select a set of independent SNPs for the relatedness analysis including the PCA used for control of ancestry. Similarly, consanguinity coefficients for probands were estimated using PC-relate, also controlling for ancestry using the first 8 PCs.

### Clinical/Diagnostic outcomes or case classification

Cases received one of either a positive (definitive positive, probable positive), inconclusive or negative case outcome based on identification of pathogenic (P), likely pathogenic (LP), variant of uncertain significance (VUS) or no primary variant(s) found (See Slavotinek et al 2023 for more details).

### Statistical analyses

The primary analysis was to compare the distribution of genetic ancestries in positive, inconclusive, and negative cases. A Kolmogorov Smirnov test was used to examine the difference in empirical cumulative distribution functions (ecdfs) of estimated genetic ancestries in positive, inconclusive, and negative cases separately for the pediatric and prenatal cases. The tests were conducted over all modes of inheritance, then stratified by mode of inheritance - Autosomal Dominant (AD), Autosomal Recessive (AR), X-linked (XL), and finally stratified by inheritance pattern (AD de novo, AD inherited, AD inheritance unknown, AR homozygous, AR compound heterozygous, XL). The significance levels were P<0.002, P<0.0007, P<.0003 for the overall, mode of inheritance and inheritance pattern stratified analyses to account for multiple testing (Bonferroni correction).

Because of non-normality and discontinuity in the genetic ancestry distributions, we created five bins of genetic ancestry as follows: 0-12.5%; >12.5-37.5%; >37.5%-62.5%; >62.5%-87.5%; >87.5-100%. These intervals were selected to reflect ranges of values around number of grandparents of differing genetic ancestry (i.e. 0, 1, 2, 3, 4). Non-parametric Cochran-Armitage trend tests were also conducted to determine whether there was a linear trend between diagnostic yield and estimated genetic ancestry (in the genetic ancestry bins). The negative cases were used as the control/reference. This was repeated in analyses stratified by mode of inheritance and inheritance pattern. To account for multiple testing, the significance levels were P<0.002, P<0.0007, P<.0003 for the overall, mode of inheritance and inheritance pattern stratified analyses.

Unpaired t-tests were used to compare means of estimated consanguinity coefficients between positive cases with various inheritance patterns vs negative cases (P-value threshold was 0.002 with Bonferroni correction, two-tailed test).

## Results

### Race/ethnicity and genetic ancestry of P^3^EGS participants

The parents of probands in the P^3^EGS cohort were racially and ethnically diverse. Among parents of pediatric probands, 40.7% were Latino(a), 18.6% White, 4.7% East Asian, 3.9% African American, 2.6% Central Asian, 2.6% South Asian, 2.3% Middle Eastern, 1.1% Native American, 0.9% Pacific Islander, 7.2% multiethnic, and 15.4% missing. Among prenatal probands’ parents, the race/ethnicity breakdown was 36.4% White, 15.5% Latino(a), 9.0% East Asian, 5.4% South Asian, 0.9% African American, 9.2% multiethnic, and 22.0% missing (Table 1).

**Table 1:** Distribution of Self-identified Race/Ethnicity of parents of pediatric and prenatal participants in the P^3^EGS cohort.

|  | **Pediatric** |  | **Prenatal** |  |
| --- | --- | --- | --- | --- |
| **Race/Ethnicity categories** | **N (% of Maternal race/ethnicity total)** | **N (% of Paternal race/ethnicity total)** | **N (% of Maternal race/ethnicity total)** | **N (% of Paternal race/ethnicity total)** |
| **Black/African American (AF)** | 21 (3.97%) | 20 (3.78%) | 3 (0.95%) | 3 (0.95%) |
| **Central Asian (CA)** | 14 (2.65%) | 14 (2.65%) | 0 (0%) | 0 (0%) |
| **East Asian (EA)** | 28 (5.29%) | 22 (4.16%) | 31 (9.81%) | 26 (8.23%) |
| **White/European American (EU)** | 96 (18.15%) | 101 (19.09%) | 115 (36.39%) | 115 (36.39%) |
| **Hispanic/Latino(a) (LT)** | 228 (43.1%) | 203 (38.37%) | 49 (15.51%) | 49 (15.51%) |
| **Middle Eastern or North African/Mediterranean (ME)** | 12 (2.27%) | 12 (2.27%) | 5 (1.58%) | 5 (1.58%) |
| **American Indian, Native American, Alaska Native (NAT)** | 6 (1.13%) | 6 (1.13%) | 0 (0%) | 0 (0%) |
| **Native Hawaiian/Pacific Islander (PI)** | 4 (0.76%) | 5 (0.95%) | 0 (0%) | 0 (0%) |
| **South Asian (SA)** | 14 (2.65%) | 13 (2.46%) | 18 (5.7%) | 16 (5.06%) |
| **AF, EU** | 6 (1.13%) | 2 (0.38%) | 2 (0.63%) | 3 (0.95%) |
| **EA, EU** | 4 (0.76%) | 2 (0.38%) | 6 (1.9%) | 1 (0.32%) |
| **EU, LT** | 9 (1.7%) | 6 (1.13%) | 8 (2.53%) | 14 (4.43%) |
| **EU, NAT** | 8 (1.51%) | 11 (2.08%) | 4 (1.27%) | 3 (0.95%) |
| **Other 2 combination R/E** | 8 (1.51%) | 8 (1.51%) | 6 (1.9%) | 2 (0.63%) |
| **3 or more R/E** | 7 (1.32%) | 5 (0.95%) | 6 (1.9%) | 3 (0.95%) |
| **Missing (Unknown/ None of these fully describe me/ Prefer not to answer)** | 64 (12.1%) | 99 (18.71%) | 63 (19.94%) | 76 (24.05%) |
| **Total** | 529 (100%) | 529 (100%) | 316 (100%) | 316 (100%) |

Results of PC analysis are given in Supplementary Figures 1-3. The first 6 PCs depict African, European, East Asian, Native American, South Asian, Middle Eastern and Pacific Islander genetic ancestries. The P^3^EGS cases reflect all these ancestries, with the largest components being European, Native American and East Asian.

The correspondence between self-identified race/ethnicity and estimated individual genetic ancestry proportions of the 1325 exome sequenced parents is visualized in Figure 1. This includes those whose race/ethnicity information was missing. As shown previously,^12^ for those reporting a single race/ethnicity there is a high correspondence between genetic ancestry and self-reported race/ethnicity (Figure 1a). For example, those reporting East Asian race/ethnicity have near 100% East Asian genetic ancestry; the same is true for those reporting South Asian, White/European, and Middle Eastern race/ethnicity. Those reporting African American race/ethnicity have admixed African and European genetic ancestry, while Latino(a) participants have primarily Native American and European genetic ancestry, with a modest contribution of African and Middle Eastern ancestry. The genetic ancestry of Central Asians appears to be intermediate between South Asian and European/Middle Eastern. The genetic ancestry distribution of those with missing race/ethnicity appears quite comparable to the overall distribution of those with information, reflecting largely European genetic ancestry, mixed European/Native American genetic ancestry, and East Asian and South Asian genetic ancestry. Parents who reported more than 1 race/ethnicity had a higher level of genetic admixture compared to those who reported only 1, and again there is a high correspondence between the self-reported race/ethnicities and genetic admixture for these participants (Figure 1b). The single exception is for those reporting Native American and European/white race/ethnicity. The majority of such participants have only European genetic ancestry, while the remainder are admixed European with a modest to moderate amount of Native American genetic ancestry.

**Figure 1:**
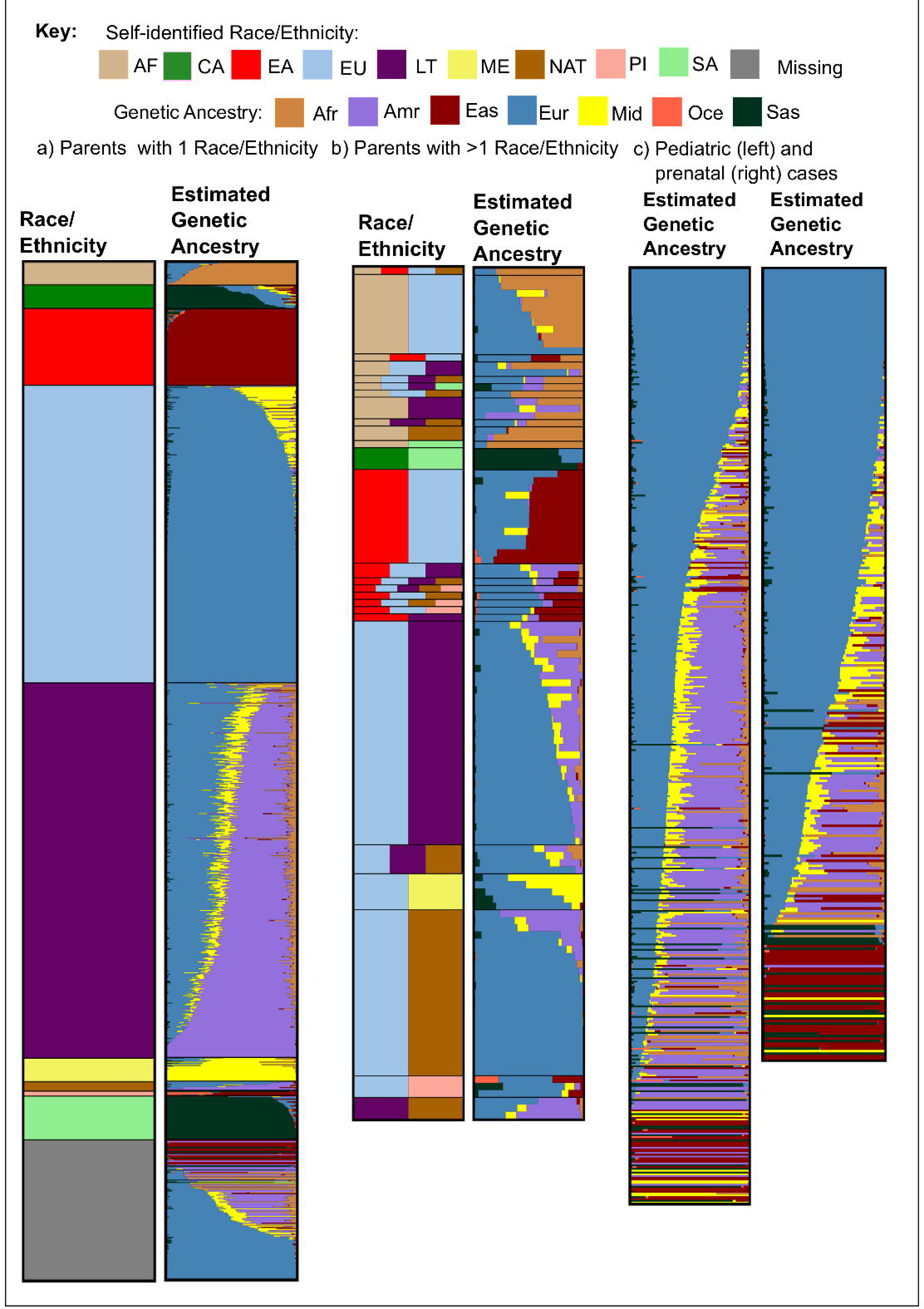
Correspondence between estimated genetic ancestry proportions and self-reported race/ethnicity of parents of P3EGS patients; and estimated genetic ancestry of pediatric and prenatal patients. Each horizontal bar in the “Estimated Genetic ancestry column” represents one parent or patient, and the “Race/Ethnicity” column corresponds to the self-reported R/E of the parent. Genetic ancestry proportions/ percentages were estimated from exome sequencing data using Admixture software with unrelated human genome diversity (HGDP) samples from gnomAD as reference samples/populations. The self-reported race/ethnicity categories are: African American or Black (AF), Central Asian (CA), East Asian (EA), White-European (EU), Latino(a) (LT), Middle Eastern (ME), Native American (NAT), Pacific Islander (PI), and South Asian (SA). The genetic ancestry groups are: Africa (Afr), Native America (Amr), East Asia (Eas), Europe (Eur), Middle East (Mid), Oceania (Oce), South Asia (Sas).

This observation is comparable to what has been reported previously.^12^ The average genetic ancestry proportions in the pediatric cases were: 41.6% European, 28.9% Native American, 7.2% East Asian, 8.6% Middle Eastern, 7.3% African, and 6.2% South Asian. The average genetic ancestry proportions in the prenatal cases were: 56.8% European, 10.6% Native American, 12.9% East Asian, 7.8% Middle Eastern, 4.5% African, and 7.2% South Asian. The average estimated Oceanian genetic ancestry was less than 1% in both pediatric and prenatal cases so we did not include in subsequent analyses.

### Genetic ancestry and diagnostic yield

The diagnostic yield was significantly higher in pediatric compared to prenatal cases.^7^ Overall, out of 529 pediatric probands, 141 (26.7%) had a positive (definitive + probable positive) case outcome and 73 (13.8%) had an inconclusive case outcome, while among 316 prenatal probands, 60 (19%) had a positive case outcome and 19 (6%) had an inconclusive case outcome.

The majority of the positive cases had an AD mode of inheritance: 70% and 65% in the pediatric and prenatal arms of the study, respectively, compared to 18% and 25%, respectively, that had AR inheritance. Compared to the positive cases, the inconclusive cases had a lower percentage that were of AD inheritance (41.1% pediatric, 60% prenatal) and a higher proportion of AR inheritance (45% pediatric, 30% prenatal).

For each of the 6 genetic ancestries, there was no statistically significant difference in genetic ancestry distributions between positive, negative, and inconclusive outcomes in both pediatric and prenatal cases (Figure 2): P-values from Kolmogorov-Smirnoff tests comparing genetic ancestry distributions in positive vs negative and inconclusive vs negative cases within each genetic ancestry group were all greater than 0.1 and not statistically significant.

**Figure 2:**
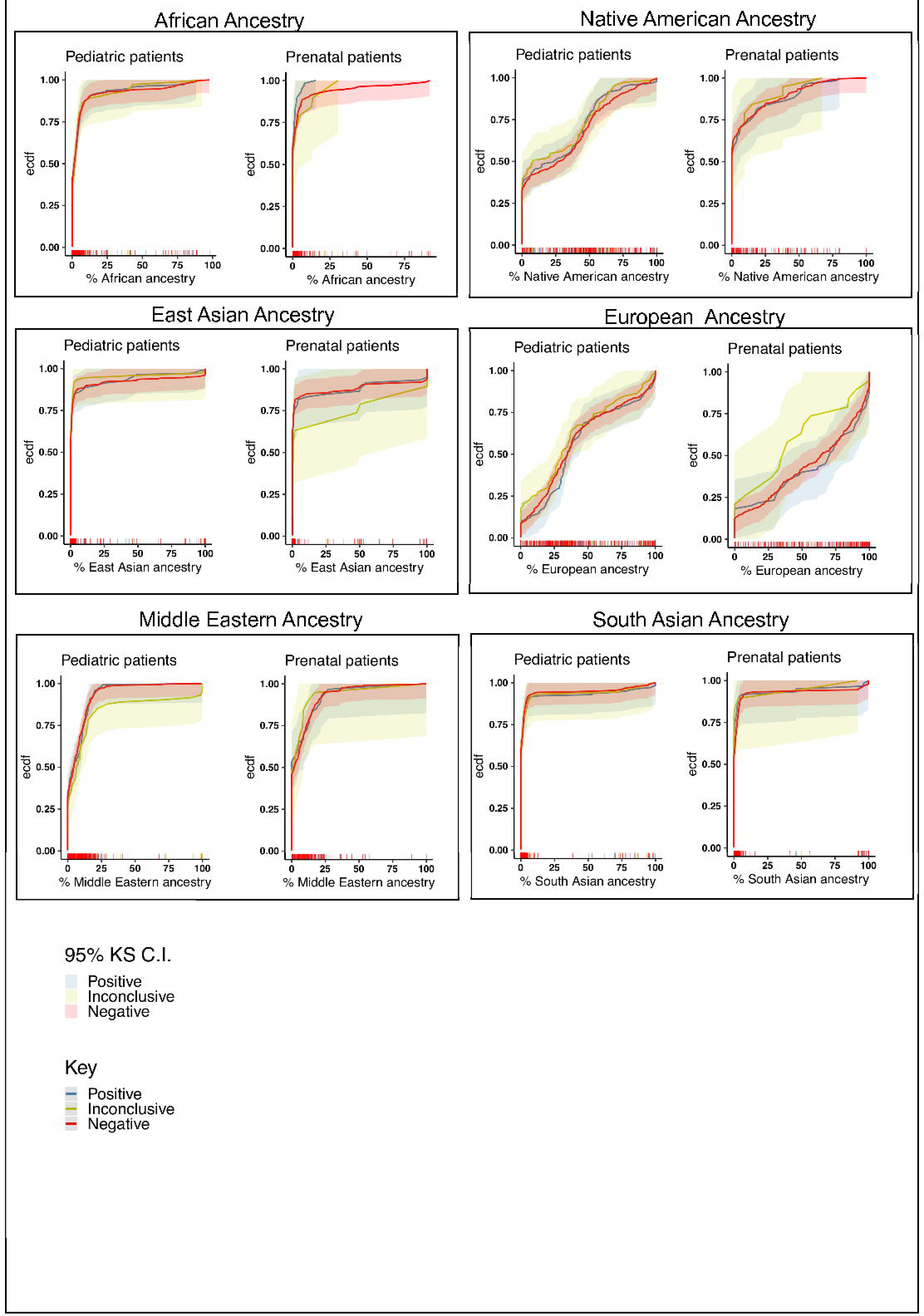
Empirical cumulative distribution functions (ecdf) and their corresponding 95% confidence interval (C.I.) bands of estimated genetic ancestries stratified by outcome — positive, inconclusive, and negative cases, and by pediatric or prenatal patients. There was no statistically significant difference between ecdfs in negative and positive cases, or negative and inconclusive cases in any of the genetic ancestries. Statistics were performed using Kolmogorov Smirnov (KS) test. The short vertical lines on the x axis represent patients, ordered by their % genetic ancestries, and colored by outcomes as seen in the legend/key. KS 95% C.I. bands for ecdfs were calculated using Dvoretzky-Kiefer-Wolfowitz (DKW) inequality.

The distribution of estimated genetic ancestries of probands was observed to be both continuous and discrete in the different genetic ancestry groups. For Amr and Eur, the estimated genetic ancestries were continuous, and in Eas, Sas, Afr, Mid, the distribution of estimated genetic ancestries were more discrete. A clear example can be seen in Figure 2, in the estimated East Asian ancestry panel, in which the groups of cases are clumped around the 0, 25, 50, 75, 100 % estimated East Asian Ancestry mark, with few cases in between. These percentages represent the number of grandparents from that ancestral population (0, 1, 2, 3 or 4). For that reason, genetic ancestry bins, containing frequency of cases (and their diagnostic yield) within estimated genetic ancestry ranges were made to best capture variation in diagnostic yield in estimated genetic ancestries (see Methods).

By the Cochran-Armitage test, there was no significant association between any genetic ancestry and diagnostic yield (Table 2). However, Mid genetic ancestry was significantly positively associated with an inconclusive outcome among pediatric probands, largely driven by the 78% (7/9) inconclusive rate in the highest Mid ancestry bin, compared to 11% (1/9) among negative cases (Table 2). This association was not observed in prenatal cases (Table 2).

**Table 2:**
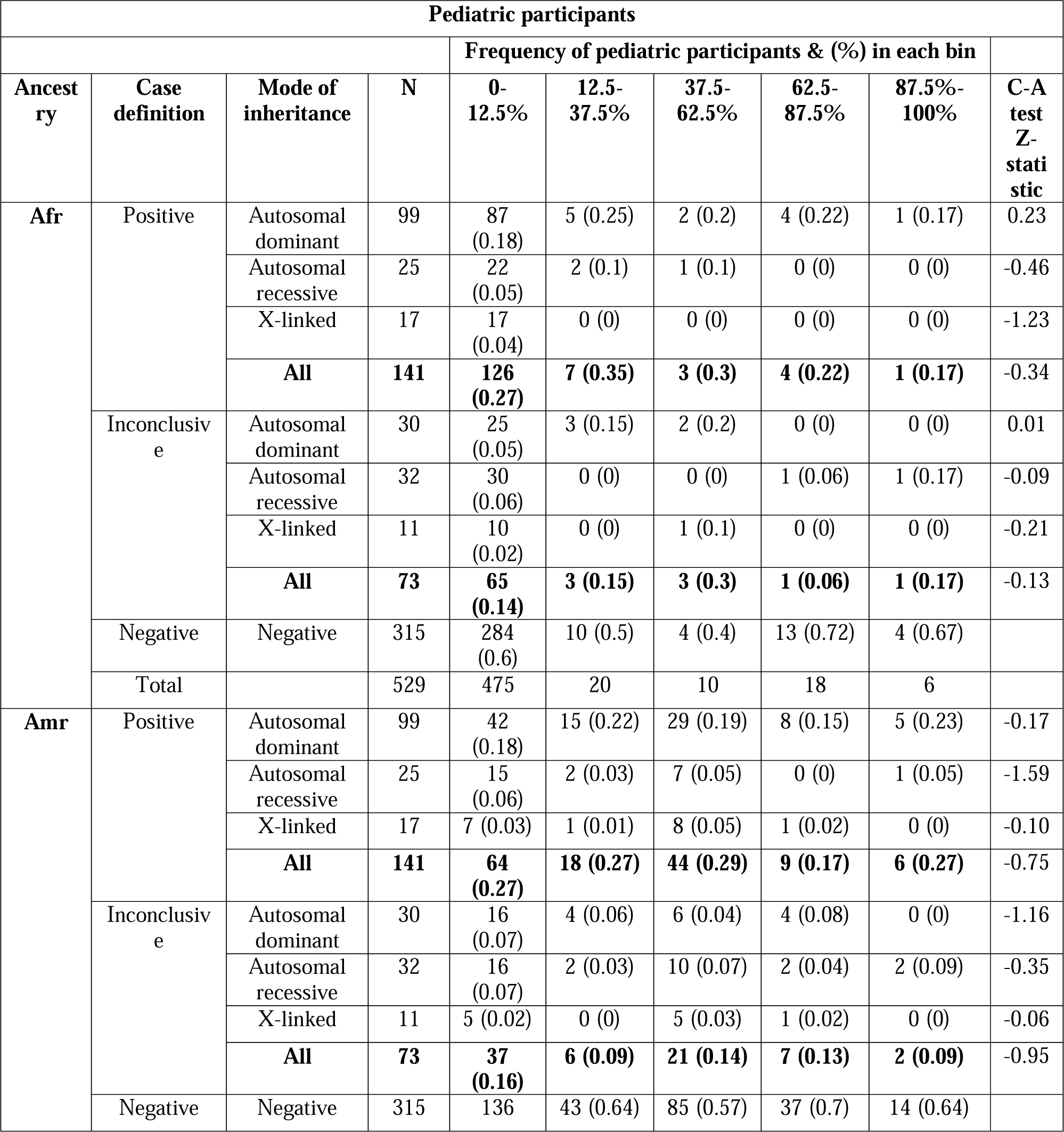
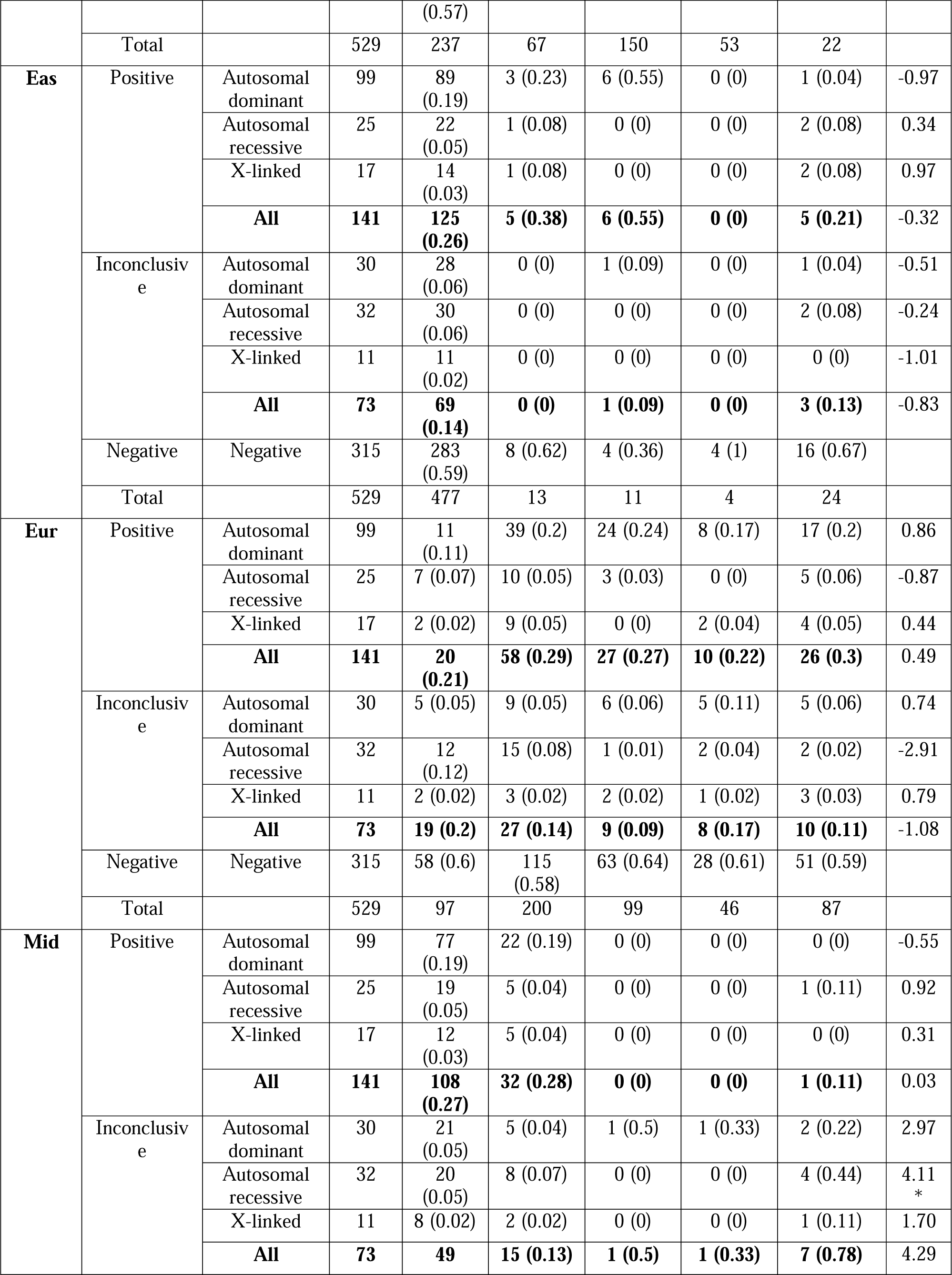
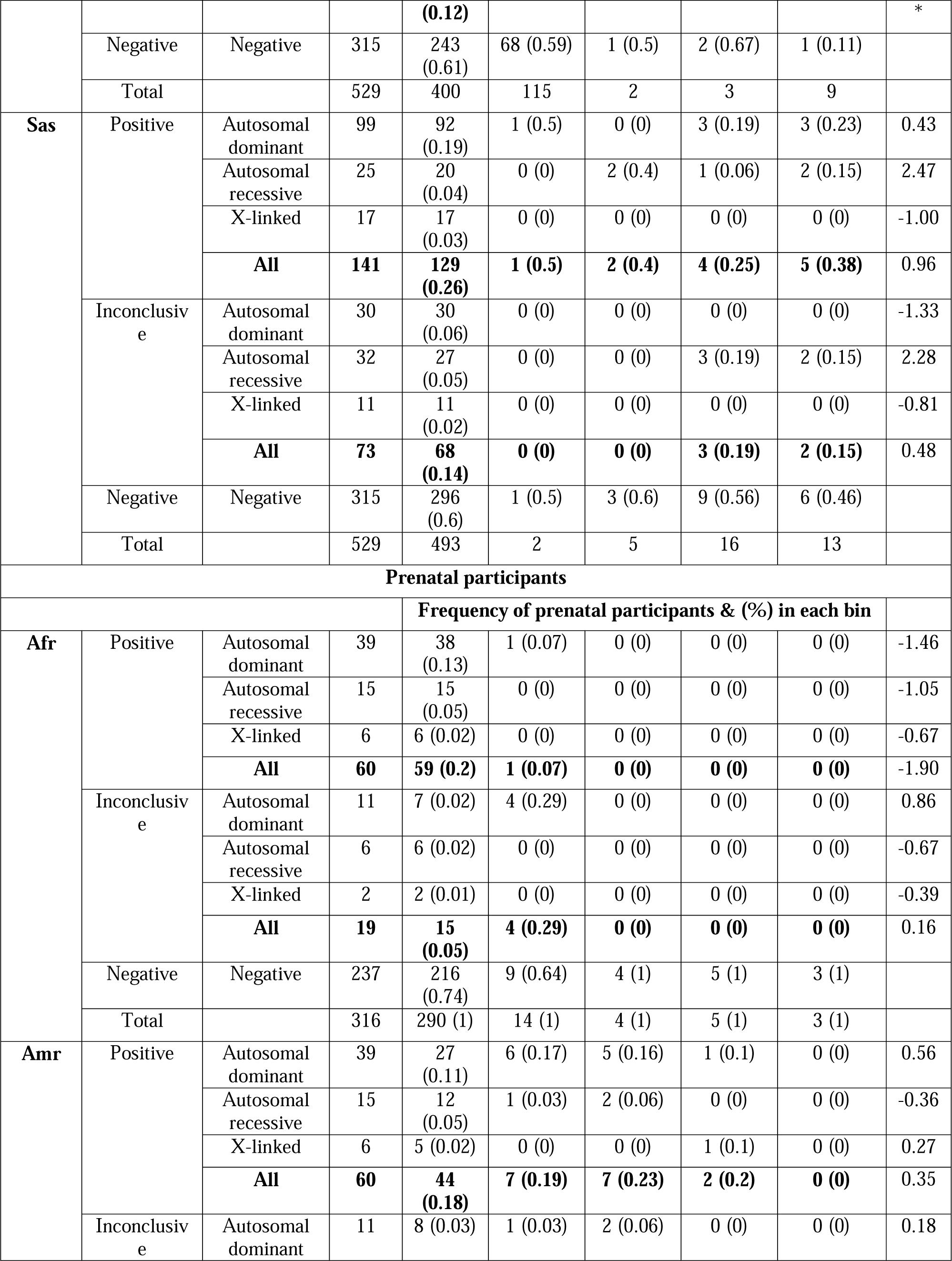
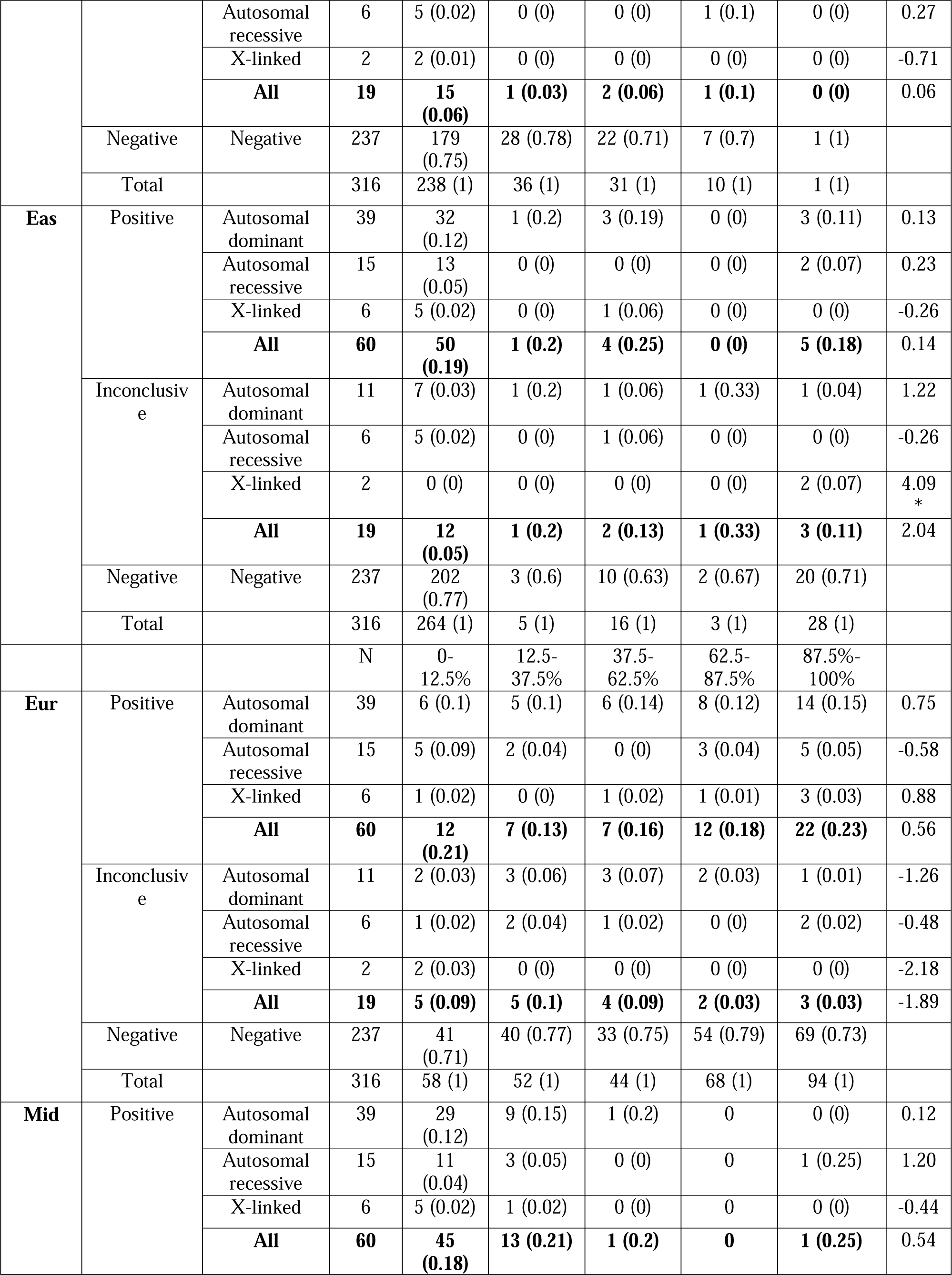
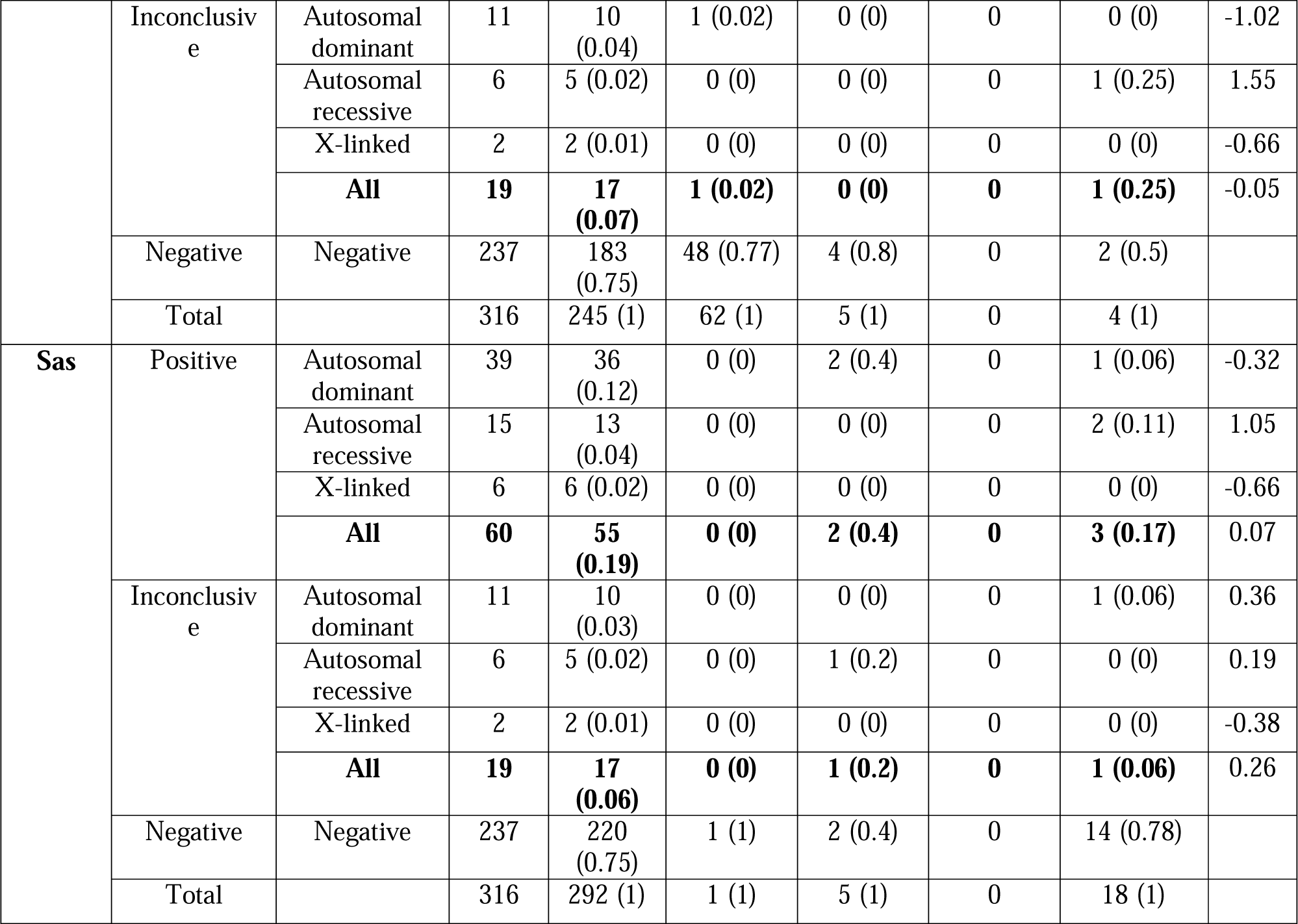
Frequency and diagnostic yield of pediatric and prenatal participants by genetic ancestry bins, stratified by mode of inheritance. Diagnostic yield (in brackets) was measured as a proportion of total number of cases in that ancestry range. Cochran-Armitage (C-A) trend test was conducted to test if diagnostic yield varies linearly with increase in estimated genetic ancestry relative to negative cases. Genetic ancestry proportions/ percentages were estimated from exome sequencing data using Admixture software with unrelated human genome diversity (HGDP) samples from gnomAD as reference samples/populations.

### Genetic ancestry and diagnostic yield stratified by mode of inheritance and inheritance patterns

Similarly, there was no statistically significant reduction in positive cases compared to negative cases associated with any estimated genetic ancestry in pediatric or prenatal cases, when the positive cases were stratified by mode of inheritance (AD, AR, XL) (Table 2). In contrast, there was a significant association of Eas genetic ancestry with XL inheritance among inconclusive prenatal cases. However, this was due entirely to 2 inconclusive cases of XL inheritance in the highest bin of Eas ancestry, whereas no such association was observed in pediatric inconclusive cases. There was also a statistically significant association between estimated Mid ancestry and AR inheritance among pediatric inconclusive cases, and a similar trend in this direction in the prenatal inconclusive cases, although numbers were quite small.

We further broke down the AR cases into homozygotes and compound heterozygotes (Supplementary Table 1, Supplementary Table 2). The association of inconclusive pediatric AR outcomes with Mid ancestry was observed only among homozygous outcomes, and not compound heterozygotes (Supplementary Table 1). Among prenatal cases, we again saw a positive association of Mid ancestry with both positive and inconclusive homozygous AR outcomes. A similar pattern was observed with Sas ancestry. In the pediatric cases, Sas ancestry was positively associated with both positive and inconclusive homozygous AR outcomes (Supplementary Table 1) but only a modest positive trend in the prenatal cases (Supplementary Table 2).

### Consanguinity coefficient and diagnostic yield

Both positive and inconclusive AR (homozygous) cases were associated with an increased consanguinity coefficient (F) among the combined pediatric and prenatal cases (Table 3). There was a statistically significant increase in mean F in AR (homozygous) outcomes among both positive and inconclusive cases compared to negative cases by unpaired t-test (P-value = .002).

**Table 3:** Mean and standard error (SE) of consanguinity coefficients (F) by Inheritance pattern in 845 P^3^EGS participants. There was a statistically significant increase in mean estimated Fs in Autosomal Recessive (homozygous) outcome among both positive and inconclusive cases compared to negative cases. Fs were estimated from exome sequencing data using PC-Relate. Comparison between means of estimated F in various inheritance patterns vs negative was done using an unpaired T-test (T-test P values threshold was 0.002, after Bonferroni correction). * Indicates P-value less than threshold.

| **Outcome** | **Inheritance pattern** | **N** | **Mean F** | **SE** | **T-test statistic (mean comparisons with negative)** | **P-value (of t-test)** |
| --- | --- | --- | --- | --- | --- | --- |
| **Positive** | **Autosomal dominant De novo** | **102** | -0.0076 | 0.0024 | -1.64 | >0.1 |
|  | **Autosomal dominant inherited** | **18** | -0.00734 | 0.0031 | -1.28 | >0.1 |
|  | **Autosomal dominant unknown** | **18** | -0.00323 | 0.0048 | -0.05 | >0.1 |
|  | **Autosomal recessive (compound heterozygous)** | **22** | -0.0148 | 0.0090 | -1.30 | >0.1 |
|  | **Autosomal recessive (homozygous)\*** | **18** | 0.043 | 0.0115 | 3.98 | 0.00092 |
|  | **X-linked** | **23** | -0.00386 | 0.0037 | -0.22 | >0.1 |
| **Negative** | **Negative** | **552** | -0.00298 | 0.0014 |  |  |
| **Inconclusive** | **Autosomal dominant De novo** | **18** | 0.000677 | 0.0080 | 0.45 | >0.1 |
|  | **Autosomal dominant inherited** | **15** | -0.0102 | 0.0068 | -1.04 | >0.1 |
|  | **Autosomal dominant unknown** | **8** | -0.00969 | 0.0058 | -1.12 | >0.1 |
|  | **Autosomal recessive (compound heterozygous)** | **11** | -0.0204 | 0.0071 | -2.41 | 0.035 |
|  | **Autosomal recessive (homozygous)\*** | **27** | 0.0494 | 0.0111 | 4.68 | 7.21E-05 |
|  | **X-linked** | **13** | 0.00925 | 0.0105 | 1.15 | >0.1 |

### Consanguinity coefficient and race/ethnicity

We examined the AR homozygous positive and inconclusive cases by self-reported race/ethnicity of the parents and consanguinity coefficient of the proband, as well as the variant frequencies. Among 18 positive cases, 9 had consanguinity coefficients greater than .01 (average of .066, minimum .037, maximum .120). For 3 of these cases the parents were South Asian, in 2 cases the parents were Central Asian, in 2 cases the parents were Middle Eastern, and in one case each the parents were Latino(a) and East Asian. Among 9 cases with consanguinity coefficients less than .01 (average of -.005), 6 had parents that were Latino(a), and one each were Central Asian, African American, and white. For all cases, the frequencies of P and LP variants estimated from gnomAD (based on the genetic ancestry estimates of the proband) were uniformly low; all were below 0.0001 except for a non-consanguineous Central Asian (.00014) and non-consanguineous African American (.00041) case. It is notable that among the 9 cases with consanguinity, 7 were Middle Eastern, Central or South Asian, while among the 9 cases without consanguinity, 1 case was Middle Eastern, Central or South Asian.

Among 26 AR homozygous inconclusive cases (with VUSs), 15 had consanguinity coefficients greater than .01 (average of .066, minimum .026, maximum .128). In this group, for 5 cases the parents were Middle Eastern, in 5 cases the parents were Latino(a), in 2 cases each the parents were Central Asian and South Asian, and in one case the parents were East Asian. In contrast, among 11 cases with consanguinity coefficients less than .01 (average of -.011), for 7 the parents were Latino(a), for 2 the parents were South Asian, and in one each the parents were white and African American. Here again it is notable that among the 15 cases with consanguinity, 9 had parents who identified as Middle Eastern, Central Asian, or South Asian, while among the 11 cases without consanguinity, the parents identified with these racial/ethnic groups for only 2. For these inconclusive AR homozygous cases, the P/LP allele frequencies were again all below .0001 in frequency except in 2 cases, one East Asian consanguineous case with allele frequency .0032, and one Latino(a) non-consanguineous case with allele frequency .00023. These variants represent ancestry-specific founder variants.

### Recurrent variants in the P^3^EGS cohort

In searching for possible founder variants, we found 4 recurrent variants in 3 different genes among 8 different cases (Supplementary Table 3). Of note, the recurrent variants were all *de novo*, and therefore do not represent founder variants.

## Discussion

Among both pediatric and prenatal cases, we observed no reduction in overall diagnostic yield (definitive+ probable positive) from ES associated with any of the estimated genetic ancestry groups (Afr, Amr, Eas, Eur, Mid, Sas). Similarly, there was no reduction or increase in the rate of inconclusive outcomes associated with any of the genetic ancestries, with the single exception of a positive association with Middle East ancestry. Of 9 pediatric cases with primarily (>87.5%) Middle East genetic ancestry, 7 (78%) had an inconclusive result, including 2 AD, 4 AR and 1 XL, compared to 12% for the rest of the cohort. There were 4 prenatal cases with majority Middle East genetic ancestry; 1 of these had an inconclusive result (AR).

The mode of inheritance distribution also differed significantly between positive and inconclusive outcomes, with a higher proportion of AD de novo results for positive versus inconclusive cases,^7^ likely a direct reflection of the ACMG criteria, for which de novo status of a variant is considered a primary criterion for pathogenicity determination. Most of our cases that were classified as inconclusive were due to variant uncertainty,^7^ and the majority of these VUSs were inherited variants or inheritance uncertain. We also observed a shift in mode of inheritance by genetic ancestry among our cases. AR homozygous inheritance was positively associated with Middle East and South Asian genetic ancestry among both positive and inconclusive pediatric and prenatal cases. We also showed that these trends were largely due to consanguinity associated with these ancestries. Thus, while the overall diagnostic yield was not diminished in any non-European genetic ancestry, the pattern of inheritance varied. And the sole positive association of the inconclusive rate with Middle East genetic ancestry was largely attributable to 5 AR homozygous cases.

Some studies have suggested that diagnostic yield from ES and other genetic tests is lower in non-white race/ethnicity groups, such as African American, or Native American ^3,13^ possibly due to underrepresentation of data from non-white populations in genetic variant databases.^2,3,14^ However, the clinical context is important in evaluating the association of race/ethnicity or genetic ancestry with diagnostic yield. For example, in genetic testing studies of hearing loss in which children underwent comprehensive genetic testing (CGT) and panel testing, Hispanic/Latino(a) and African American children were less likely to have a definitive genetic diagnosis compared to white or Asian children.^5,15^ This was due to the fact that likely causal variants in the African American and Latino(a) children had not yet been documented in prior studies (and therefore also not appearing on variant-specific panels), in contrast to some of the more common causal variants found in white and Asian children. When the authors reduced the ACMG criterion of prior association with disease and solely used in silico functional prediction, there was no difference in diagnostic yield. It appears that the requirement for prior evidence regarding a specific variant (as opposed to predicted functional evidence) can have a significant impact on diagnostic yield; an example from newborn screening demonstrated a reduction of diagnostic yield from 88% to 55% when requiring prior curation of a variant as P/LP as opposed to functional prediction with no prior curation, yet without a dramatic effect on false positive rate (increase from 0.6% to 1.6%).^16^ It is therefore important to consider the role of prior evidence of pathogenicity or likely pathogenicity for a variant in assessing genetic ancestry influences on diagnostic yield.

In our study, cases were selected with a broad range of clinical phenotypes, with no prior assumptions about potential mode of inheritance. The majority of our positively diagnosed pediatric and prenatal cases were due to P/LP variants in AD genes (69%), and the majority of the variants were de novo (74% confirmed but possibly as high as 87% due to inheritance uncertainty). All of our XL cases were also dominant, and the majority arose de novo.^7^ By contrast, 9% of the positive cases were due to inherited AD variants, and 20% had AR variants, nearly all of which were inherited. As expected, we observed no genetic ancestry association with de novo variants as these presumably occur independently from an individual’s genetic ancestry. However, we also saw no genetic ancestry associations in the inherited AD or AR cases. This was largely a reflection that nearly all variants were quite rare (frequency < .0001), and, with few exceptions, likely did not reflect founder mutations in any of the conditions or groups studied. The possible exceptions are variants observed in AR homozygous cases with low consanguinity coefficients as well as AR compound heterozygotes. Of note, we found no genetic ancestry associations or even trends for the pediatric or prenatal AR compound heterozygous cases. On the other hand, we did observe an excess of Native American genetic ancestry among 9 AR homozygotes without consanguinity, reflecting that 6 of the 9 had parents who self-reported Latino(a) race/ethnicity, and suggesting the possibility of founder variants in some American indigenous populations.

Among the inconclusive cases, the proportion of inherited cases is substantially higher at 58% (53/92). Yet here also, we found no association with any of the genetic ancestries tested for the inherited AD and compound heterozygous AR cases. Again, this suggests that while variant uncertainty may have led to this collection of outcomes, there was no bias towards non-European ancestries, likely because of the lack of elevated frequency of founder variants underlying the disorders identified. In the entire cohort, we identified only one P/LP/VUS variant with increased frequency – the AR VUS c.636G>C (p.[Gln212His]) (rs201590882) in *ARMC9* in an East Asian case (gnomAD frequency of .003 in East Asians). Furthermore, the four variants found twice among our cases were all de novo and not inherited.

The increased AR homozygous inheritance cases in high Mid and Sas ancestry pediatric patients, corresponding with statistically increased estimated consanguinity coefficients, was expected. It is well documented that certain population groups such as those from the Middle East, and South and Central Asia, have increased F, which increases autozygosity and hence the rate of AR homozygous cases.^17,18^

We also note that our results on ancestry are a direct reflection of the clinical setting of rare, undiagnosed diseases and implementation of the ACMG criteria for variant annotation. Those criteria place a special emphasis on de novo inheritance, leading to a higher proportion of de novo AD variants in positive cases compared to inconclusive cases in our study. While there was a lack of founder variants underlying the genetic etiology of the cases in our study, this phenomenon may not be general – for example in the study of known predominantly AR diseases (such as hearing loss or inborn errors of metabolism), where ancestry associations may still be present depending on variant annotation requirements. Thus, our results should not necessarily be considered representative of all clinical testing scenarios.

In summary, in this ancestrally diverse cohort of pediatric and prenatal cases with different clinical indications, there was no reduction in diagnostic yield associated with any genetic ancestry group. Consanguinity may increase the relative proportion of cases with AR homozygous inheritance among those with Middle East and South Asian genetic ancestry but did not alter the overall diagnostic yield, although our number of cases with these ancestries was modest. This empirical study improves our understanding and provides support for the ethical and equitable use of exome sequencing in diagnosis of previously undiagnosed but potentially Mendelian disorders across all ancestral populations.

## Data Availability

Sequencing data has been uploaded to the Analysis Visualization and Informatics Lab-space
(AnVIL) at the National Human Genome Research Institute. Clinical data is available from the
authors on reasonable request.

## Data Availability

Sequencing data has been uploaded to the Analysis Visualization and Informatics Lab-space (AnVIL) at the National Human Genome Research Institute. Clinical data is available from the authors on reasonable request.

### Acknowledgements

Research reported in this publication was supported by the National Human Genome Research Institute at the National Institutes of Health under Award Number U01HG009599. The content is solely the responsibility of the authors and does not necessarily represent the official views of the National Institutes of Health.

## Author Contributions

*Conceptualization* – Yusuph Mavura, Neil Risch, Anne Slavotinek, Barbara Koenig, Sara Ackerman, Pui-Yan Kwok, Mary E, Norton

*Data Curation* - Jessica van Ziffle, W. Patrick Devine, Ugur Hodoglugil, Pierre-Marie Martin, Mark Kvale

*Formal Analysis* - Yusuph Mavura, Neil Risch

*Funding acquisition* - Anne Slavotinek, Barbara Koenig, Sara Ackerman, Neil Risch, Pui-Yan Kwok, Mary E. Norton

*Project Administration* - Nuriye Sahin-Hodoglugil

*Investigation* – Yusuph Mavura, Neil Risch

*Writing - original draft* – Yusuph Mavura, Neil Risch

*Writing - review and editing* - all authors

## Ethics Declaration

The study was approved by the UCSF Institutional Review Board (IRB) (protocols 17-22504 and 17-22420), the Fresno Community Medical Center IRB (protocol 2019024), and was registered as two clinical trials (“Clinical Utility of Pediatric Whole Exome Sequencing”, NCT03525431 and “Clinical Utility of Prenatal Whole Exome Sequencing”, NCT03482141). Written informed consent was provided by adult participants >18 years of age, or by parents or legal guardians on behalf of their children <18 years of age or >18 years of age who were unable to consent independently. Assent was obtained from minors and intellectually disabled adults whenever possible.

