## Supplementary Material for "Diagnostic Yield of Exome Sequencing in a Diverse Pediatric and Prenatal Population is not Associated with Genetic Ancestry"

**Categorization of Race/Ethnicity for Parents of P^3^EGS Probands**

In terms of race/ethnicity/nationality, the P^3^EGS parents were asked to respond to all categories that best describe them among: a) American Indian, Native American or Alaska Native, b) Asian-Filipino, c) Asian-Central/South Asian (Indian, Pakistani, Afghani), d) Asian-Vietnamese, e) Asian-Hmong, f) Asian-Korean, g) Asian-Japanese, h) Asian-other (specified through free text), i) Black or African American, j) Native Hawaiian, k) Samoan, l) Other Pacific Islander (specified through free text), m) white or European American, n) Middle Eastern or North African/Mediterranean, o) Hispanic/Latino(a) – Mexican, Mexican American, Chicano/a, p) Hispanic/Latino(a) – Central American -Guatemala, El Salvador, etc., q) Hispanic/Latino(a) – South American -Peru, Chile, etc., r) Hispanic/Latino(a) – Caribbean -Puerto Rico, Cuba, etc., s) Hispanic/Latino(a) – another Hispanic or Latino origin (specified by free text), t) Prefer not to answer u) Unknown/none of these fully describe them. They also responded to the open-ended questions “What is your ancestry or ethnic origin?” and “What country were you born in?”

Based on the parental responses to the demographic questionnaires, we derived the following categories (based primarily on the selected pre-listed categories above, and further resolved using the open-ended questions): Native American (NAT) — based on category a); Latino(a) (LT) — based on categories o) to s) which were rolled up; White-European (EU) — based on category m); African American or Black (AF) — based on category i); East Asian (EA) — based on categories b), d) to g) which were rolled up; South Asian (SA) and Central Asian (CA) — by separating category c) into SA and CA based on information from the open-ended questions on ancestry and country of origin; Middle Eastern (ME) — based on category n) and Pacific Islander (PI) — based on categories j) to l) which were rolled up. The open-ended questions were also used to resolve category h) into EA, SA, or CA. Each of the parents was placed in one or more of the categories or “missing” if no information was provided.

We only included self-reported race/ethnicity categories for parents, as no self-report information is available for children or fetuses, and parents did not assign race/ethnicity categories to their offspring.

**Exome Sequencing, Quality Control and Selection of Markers for Genetic Ancestry Analyses**

Exon regions were targeted using the xGen Whole Exome Panel kit from Integrated DNA Technologies. The targeted regions were sequenced using the Illumina HiSeq 2500 sequencing system (v3 chemistry) with 100 bp paired end reads in rapid run mode. The DNA sequences were aligned to the reference published human genome GRch37 (See Slavotinek et. al 2023 for full methods ).

Variants in all VCF files with sequencing depth at or below 10 (DP<=10), and genotype quality equal to or less than 20 (GQ <=20) were filtered out using GATK. ^19^ The VCF files were then lifted over from human genome reference version GRCh37 to GRCh38 using the Picard tool in GATK suite of tools. ^19^ Human Genome Diversity Panel (HGDP) whole genome sequencing samples from the GnomAD V3 call set ^20^ ^21^ were used as the reference for genetic ancestry and admixture estimation (N=829 unrelated individuals). The HGDP samples were all mapped to the GRCh38 reference sequence.

High-performance markers were selected from the HGDP and P^3^EGS data for downstream genetic ancestry, admixture, relatedness, and consanguinity analysis using the following criteria:

1) Restriction of markers in the HGDP dataset to exome sequenced regions in the P^3^EGS dataset. This was conducted using bcftools. ^22^

2) MAF >= 0.05 in any of 7 supergroups in GnomAD HGDP unrelated individuals:

i) African (ii) Native American, iii) South Asian iv) East Asian, v) European, vi) Middle Eastern, vii) Oceanian.

3) Only biallelic, autosomal SNPs, with a call rate > 95% in exome regions that were sequenced were selected (This was done in both HGDP and P^3^EGS cohorts separately). The resulting markers in the HGDP cohort (N=105,956) were intersected with markers from the P^3^EGS cohort sample VCFs, resulting in N=95,173 markers. Variants in regions known to affect principal components (PCs) (HLA region on chromosome 6p, inversion on chromosome 8p23 and inversion on chr 17q21, GRCh38 build) were removed resulting in 82,349 markers after filtering. After linkage disequilibrium pruning (0.5 kb in a 5000 kb window), 53,665 high-performance markers for principal components analysis and genetic admixture analysis remained.
