## Supplementary Tables for "Diagnostic Yield of Exome Sequencing in a Diverse Pediatric and Prenatal Population is not Associated with Genetic Ancestry"

**Supplementary Table 1: Frequency of pediatric subjects by genetic ancestry bins, stratified by extended inheritance pattern.**

|  |  |  | | **Frequency of pediatric subjects & (%) in each bin** | | | | |  |
| --- | --- | --- | --- | --- | --- | --- | --- | --- | --- |
| **Ancestry** | **Case definition** | **Inheritance Pattern** | **N** | **0-12.5%** | **12.5-37.5%** | **37.5-62.5%** | **62.5-87.5%** | **87.5-100%** | **C-A test Z-statistic** |
| **Afr** | Positive | Autosomal dominant De novo | **68** | 62 | 2 | 1 | 2 | 1 | -0.25 |
|  |  | Autosomal dominant inherited | **14** | 12 | 1 | 0 | 1 | 0 | 0.25 |
|  |  | Autosomal dominant unknown | **17** | 13 | 2 | 1 | 1 | 0 | 0.93 |
|  |  | Autosomal recessive (compound heterozygous) | **11** | 9 | 1 | 1 | 0 | 0 | 0.17 |
|  |  | Autosomal recessive (homozygous) | **14** | 13 | 1 | 0 | 0 | 0 | 0.94 |
|  |  | X-linked | **17** | 17 | 0 | 0 | 0 | 0 | -1.23 |
|  | Inconclusive | Autosomal dominant De novo | **12** | 11 | 1 | 0 | 0 | 0 | -0.66 |
|  |  | Autosomal dominant inherited | **11** | 8 | 2 | 1 | 0 | 0 | 0.56 |
|  |  | Autosomal dominant unknown | **7** | 6 | 0 | 1 | 0 | 0 | 0.18 |
|  |  | Autosomal recessive (compound heterozygous) | **8** | 7 | 0 | 0 | 0 | 1 | 0.94 |
|  |  | Autosomal recessive (homozygous) | **24** | 23 | 0 | 0 | 1 | 0 | -0.66 |
|  |  | X-linked | **11** | 10 | 0 | 1 | 0 | 0 | -0.21 |
|  | Negative | Negative | **315** | 284 | 10 | 4 | 13 | 4 |  |
| **Amr** | Positive | Autosomal dominant De novo | **68** | 28 | 9 | 21 | 6 | 4 | 0.26 |
|  |  | Autosomal dominant inherited | **14** | 8 | 2 | 3 | 1 | 0 | -1.25 |
|  |  | Autosomal dominant unknown | **17** | 6 | 4 | 5 | 1 | 1 | 0.09 |
|  |  | Autosomal recessive (compound heterozygous) | **11** | 6 | 2 | 3 | 0 | 0 | -1.27 |
|  |  | Autosomal recessive (homozygous) | **14** | 9 | 0 | 4 | 0 | 1 | -1.03 |
|  |  | X-linked | **17** | 7 | 1 | 8 | 1 | 0 | -0.10 |
|  | Inconclusive | Autosomal dominant De novo | **12** | 7 | 1 | 2 | 2 | 0 | -0.80 |
|  |  | Autosomal dominant inherited | **11** | 5 | 1 | 3 | 2 | 0 | -0.06 |
|  |  | Autosomal dominant unknown | **7** | 4 | 2 | 1 | 0 | 0 | -1.35 |
|  |  | Autosomal recessive (compound heterozygous) | **8** | 4 | 1 | 3 | 0 | 0 | -0.75 |
|  |  | Autosomal recessive (homozygous) | **24** | 12 | 1 | 7 | 2 | 2 | 0.01 |
|  |  | X-linked | **11** | 5 | 0 | 5 | 1 | 0 | -0.06 |
|  | Negative | Negative | **315** | 136 | 43 | 85 | 37 | 14 |  |
| **Eas** | Positive | Autosomal dominant De novo | **68** | 61 | 2 | 5 | 0 | 0 | -0.96 |
|  |  | Autosomal dominant inherited | **14** | 14 | 0 | 0 | 0 | 0 | -1.14 |
|  |  | Autosomal dominant unknown | **17** | 14 | 1 | 1 | 0 | 1 | 0.50 |
|  |  | Autosomal recessive (compound heterozygous) | **11** | 9 | 1 | 0 | 0 | 1 | 0.55 |
|  |  | Autosomal recessive (homozygous) | **14** | 13 | 0 | 0 | 0 | 1 | -0.02 |
|  |  | X-linked | **17** | 14 | 1 | 0 | 0 | 2 | 0.97 |
|  | Inconclusive | Autosomal dominant De novo | **12** | 12 | 0 | 0 | 0 | 0 | -1.05 |
|  |  | Autosomal dominant inherited | **11** | 9 | 0 | 1 | 0 | 1 | 0.85 |
|  |  | Autosomal dominant unknown | **7** | 7 | 0 | 0 | 0 | 0 | -0.81 |
|  |  | Autosomal recessive (compound heterozygous) | **8** | 7 | 0 | 0 | 0 | 1 | 0.60 |
|  |  | Autosomal recessive (homozygous) | **24** | 23 | 0 | 0 | 0 | 1 | -0.62 |
|  |  | X-linked | **11** | 11 | 0 | 0 | 0 | 0 | -1.01 |
|  | Negative | Negative | **315** | 283 | 8 | 4 | 4 | 16 |  |
| **Eur** | Positive | Autosomal dominant De novo | **68** | 9 | 28 | 15 | 5 | 11 | 0.24 |
|  |  | Autosomal dominant inherited | **14** | 0 | 5 | 3 | 2 | 4 | 1.88 |
|  |  | Autosomal dominant unknown | **17** | 2 | 6 | 6 | 1 | 2 | 0.08 |
|  |  | Autosomal recessive (compound heterozygous) | **11** | 1 | 4 | 2 | 0 | 4 | 1.24 |
|  |  | Autosomal recessive (homozygous) | **14** | 6 | 6 | 1 | 0 | 1 | -2.28 |
|  |  | X-linked | **17** | 2 | 9 | 0 | 2 | 4 | 0.44 |
|  | Inconclusive | Autosomal dominant De novo | **12** | 2 | 4 | 1 | 3 | 2 | 0.61 |
|  |  | Autosomal dominant inherited | **11** | 3 | 4 | 2 | 1 | 1 | -0.78 |
|  |  | Autosomal dominant unknown | **7** | 0 | 1 | 3 | 1 | 2 | 1.77 |
|  |  | Autosomal recessive (compound heterozygous) | **8** | 2 | 3 | 1 | 1 | 1 | -0.38 |
|  |  | Autosomal recessive (homozygous) | **24** | 10 | 12 | 0 | 1 | 1 | -3.19 |
|  |  | X-linked | **11** | 2 | 3 | 2 | 1 | 3 | 0.79 |
|  | Negative | Negative | **315** | 58 | 115 | 63 | 28 | 51 |  |
| **Mid** | Positive | Autosomal dominant De novo | **68** | 52 | 16 | **0** | **0** | **0** | -0.28 |
|  |  | Autosomal dominant inherited | **14** | 11 | 3 | 0 | 0 | 0 | -0.28 |
|  |  | Autosomal dominant unknown | **17** | 14 | 3 | 0 | 0 | 0 | -0.60 |
|  |  | Autosomal recessive (compound heterozygous) | **11** | 8 | 3 | 0 | 0 | 0 | 0.12 |
|  |  | Autosomal recessive (homozygous) | **14** | 11 | 2 | 0 | 0 | 1 | 1.15 |
|  |  | X-linked | **17** | 12 | 5 | 0 | 0 | 0 | 0.31 |
|  | Inconclusive | Autosomal dominant De novo | **12** | 7 | 3 | 0 | 0 | 2 | 3.79 |
|  |  | Autosomal dominant inherited | **11** | 9 | 1 | 0 | 1 | 0 | 0.66 |
|  |  | Autosomal dominant unknown | **7** | 5 | 1 | 1 | 0 | 0 | 0.87 |
|  |  | Autosomal recessive (compound heterozygous) | **8** | 5 | 3 | 0 | 0 | 0 | 0.65 |
|  |  | Autosomal recessive (homozygous) | **24** | 15 | 5 | 0 | 0 | 4 | 4.49 |
|  |  | X-linked | **11** | 8 | 2 | 0 | 0 | 1 | 1.70 |
|  | Negative | Negative | **315** | 243 | 68 | 1 | 2 | 1 |  |
| **Sas** | Positive | Autosomal dominant De novo | **68** | 61 | 1 | 0 | 3 | 3 | 1.29 |
|  |  | Autosomal dominant inherited | **14** | 14 | 0 | 0 | 0 | 0 | -0.91 |
|  |  | Autosomal dominant unknown | **17** | 17 | 0 | 0 | 0 | 0 | -1.00 |
|  |  | Autosomal recessive (compound heterozygous) | **11** | 11 | 0 | 0 | 0 | 0 | -0.81 |
|  |  | Autosomal recessive (homozygous) | **14** | 9 | 0 | 2 | 1 | 2 | 3.95 |
|  |  | X-linked | **17** | 17 | 0 | 0 | 0 | 0 | -1.00 |
|  | Inconclusive | Autosomal dominant De novo | **12** | 12 | 0 | 0 | 0 | 0 | -0.84 |
|  |  | Autosomal dominant inherited | **11** | 11 | 0 | 0 | 0 | 0 | -0.81 |
|  |  | Autosomal dominant unknown | **7** | 7 | 0 | 0 | 0 | 0 | -0.65 |
|  |  | Autosomal recessive (compound heterozygous) | **8** | 8 | 0 | 0 | 0 | 0 | -0.69 |
|  |  | Autosomal recessive (homozygous) | **24** | 19 | 0 | 0 | 3 | 2 | 2.99 |
|  |  | X-linked | **11** | 11 | 0 | 0 | 0 | 0 | -0.81 |
|  | Negative | Negative | **315** | 296 | 1 | 3 | 9 | 6 |  |

**Supplementary Table 2: Frequency of prenatal subjects by genetic ancestry bins, stratified by extended inheritance pattern.**

|  |  |  | | **Frequency of prenatal subjects & (%) in each bin** | | | | |  |
| --- | --- | --- | --- | --- | --- | --- | --- | --- | --- |
| **Ancestry** | **Case definition** | **Inheritance Pattern** | N | **0-12.5%** | **12.5-37.5%** | **37.5-62.5%** | **62.5-87.5%** | **87.5-100%** | **C-A test Z-statistic** |
| **Afr** | Positive | Autosomal dominant De novo | **34** | 33 | 1 | 0 | 0 | 0 | -1.33 |
|  |  | Autosomal dominant inherited | **4** | 4 | 0 | 0 | 0 | 0 | -0.55 |
|  |  | Autosomal dominant unknown | **1** | 1 | 0 | 0 | 0 | 0 | -0.27 |
|  |  | Autosomal recessive (compound heterozygous) | **11** | 11 | 0 | 0 | 0 | 0 | -0.90 |
|  |  | Autosomal recessive (homozygous) | **4** | 4 | 0 | 0 | 0 | 0 | -0.55 |
|  |  | X-linked | **6** | 6 | 0 | 0 | 0 | 0 | -0.67 |
|  | Inconclusive | Autosomal dominant De novo | **6** | 3 | 3 | 0 | 0 | 0 | 1.12 |
|  |  | Autosomal dominant inherited | **4** | 3 | 1 | 0 | 0 | 0 | 0.19 |
|  |  | Autosomal dominant unknown | **1** | 1 | 0 | 0 | 0 | 0 | -0.27 |
|  |  | Autosomal recessive (compound heterozygous) | **3** | 3 | 0 | 0 | 0 | 0 | 0.19 |
|  |  | Autosomal recessive (homozygous) | **3** | 3 | 0 | 0 | 0 | 0 | 0.19 |
|  |  | X-linked | **2** | 2 | 0 | 0 | 0 | 0 | -0.39 |
|  | Negative | Negative | **237** | 216 | 9 | 4 | 5 | 3 |  |
| **Amr** | Positive | Autosomal dominant De novo | **34** | 24 | 5 | 5 | 0 | 0 | 0.22 |
|  |  | Autosomal dominant inherited | **4** | 2 | 1 | 0 | 1 | 0 | 1.43 |
|  |  | Autosomal dominant unknown | **1** | 1 | 0 | 0 | 0 | 0 | -0.51 |
|  |  | Autosomal recessive (compound heterozygous) | **11** | 10 | 1 | 0 | 0 | 0 | -1.30 |
|  |  | Autosomal recessive (homozygous) | **4** | 2 | 0 | 2 | 0 | 0 | 1.43 |
|  |  | X-linked | **6** | 5 | 0 | 0 | 1 | 0 | 0.27 |
|  | Inconclusive | Autosomal dominant De novo | **6** | 5 | 0 | 1 | 0 | 0 | -0.23 |
|  |  | Autosomal dominant inherited | **4** | 3 | 1 | 0 | 0 | 0 | -0.39 |
|  |  | Autosomal dominant unknown | **1** | 0 | 0 | 1 | 0 | 0 | 1.95 |
|  |  | Autosomal recessive (compound heterozygous) | **3** | 3 | 0 | 0 | 0 | 0 | -0.87 |
|  |  | Autosomal recessive (homozygous) | **3** | 2 | 0 | 0 | 1 | 0 | 1.24 |
|  |  | X-linked | **2** | 2 | 0 | 0 | 0 | 0 | -0.71 |
|  | Negative | Negative | **237** | 179 | 28 | 22 | 7 | 1 |  |
| **Eas** | Positive | Autosomal dominant De novo | **34** | 27 | 1 | 3 | 0 | 3 | 0.45 |
|  |  | Autosomal dominant inherited | **4** | 4 | 0 | 0 | 0 | 0 | -0.78 |
|  |  | Autosomal dominant unknown | **1** | 1 | 0 | 0 | 0 | 0 | -0.39 |
|  |  | Autosomal recessive (compound heterozygous) | **11** | 9 | 0 | 0 | 0 | 2 | 0.72 |
|  |  | Autosomal recessive (homozygous) | **4** | 4 | 0 | 0 | 0 | 0 | -0.78 |
|  |  | X-linked | **6** | 5 | 0 | 1 | 0 | 0 | -0.26 |
|  | Inconclusive | Autosomal dominant De novo | **6** | 2 | 1 | 1 | 1 | 1 | 2.42 |
|  |  | Autosomal dominant inherited | **4** | 4 | 0 | 0 | 0 | 0 | -0.78 |
|  |  | Autosomal dominant unknown | **1** | 1 | 0 | 0 | 0 | 0 | -0.39 |
|  |  | Autosomal recessive (compound heterozygous) | **3** | 2 | 0 | 1 | 0 | 0 | 0.30 |
|  |  | Autosomal recessive (homozygous) | **3** | 3 | 0 | 0 | 0 | 0 | -0.67 |
|  |  | X-linked | **2** | 0 | 0 | 0 | 0 | 2 | 4.09 |
|  | Negative | Negative | **237** | 202 | 3 | 10 | 2 | 20 |  |
| **Eur** | Positive | Autosomal dominant De novo | **34** | 6 | 4 | 6 | 7 | 11 | 0.32 |
|  |  | Autosomal dominant inherited | **4** | 0 | 1 | 0 | 1 | 2 | 0.95 |
|  |  | Autosomal dominant unknown | **1** | 0 | 0 | 0 | 0 | 1 | 1.16 |
|  |  | Autosomal recessive (compound heterozygous) | **11** | 3 | 0 | 0 | 3 | 5 | 0.75 |
|  |  | Autosomal recessive (homozygous) | **4** | 2 | 2 | 0 | 0 | 0 | -2.41 |
|  |  | X-linked | **6** | 1 | 0 | 1 | 1 | 3 | 0.88 |
|  | Inconclusive | Autosomal dominant De novo | **6** | 1 | 3 | 2 | 0 | 0 | -1.86 |
|  |  | Autosomal dominant inherited | **4** | 1 | 0 | 0 | 2 | 1 | 0.28 |
|  |  | Autosomal dominant unknown | **1** | 0 | 0 | 1 | 0 | 0 | -0.20 |
|  |  | Autosomal recessive (compound heterozygous) | **3** | 0 | 1 | 0 | 0 | 2 | 0.82 |
|  |  | Autosomal recessive (homozygous) | **3** | 1 | 1 | 1 | 0 | 0 | -1.51 |
|  |  | X-linked | **2** | 2 | 0 | 0 | 0 | 0 | -2.18 |
|  | Negative | Negative | **237** | 41 | 40 | 33 | 54 | 69 |  |
| **Mid** | Positive | Autosomal dominant De novo | **34** | 25 | 8 | 1 | 0 | 0 | 0.23 |
|  |  | Autosomal dominant inherited | **4** | 3 | 1 | 0 | 0 | 0 | -0.07 |
|  |  | Autosomal dominant unknown | **1** | 1 | 0 | 0 | 0 | 0 | -0.47 |
|  |  | Autosomal recessive (compound heterozygous) | **11** | 9 | 2 | 0 | 0 | 0 | -0.50 |
|  |  | Autosomal recessive (homozygous) | **4** | 2 | 1 | 0 | 0 | 1 | 3.12 |
|  |  | X-linked | **6** | 5 | 1 | 0 | 0 | 0 | -0.44 |
|  | Inconclusive | Autosomal dominant De novo | **6** | 6 | 0 | 0 | 0 | 0 | -1.14 |
|  |  | Autosomal dominant inherited | **4** | 4 | 0 | 0 | 0 | 0 | -0.94 |
|  |  | Autosomal dominant unknown | **1** | 0 | 1 | 0 | 0 | 0 | 1.26 |
|  |  | Autosomal recessive (compound heterozygous) | **3** | 3 | 0 | 0 | 0 | 0 | -0.81 |
|  |  | Autosomal recessive (homozygous) | **3** | 2 | 0 | 0 | 0 | 1 | 2.94 |
|  |  | X-linked | **2** | 2 | 0 | 0 | 0 | 0 | -0.66 |
|  | Negative | Negative | **237** | 183 | 48 | 4 | 0 | 2 |  |
| **Sas** | Positive | Autosomal dominant De novo | **34** | 31 | 0 | 2 | 0 | 1 | -0.13 |
|  |  | Autosomal dominant inherited | **4** | 4 | 0 | 0 | 0 | 0 | -0.54 |
|  |  | Autosomal dominant unknown | **1** | 1 | 0 | 0 | 0 | 0 | -0.27 |
|  |  | Autosomal recessive (compound heterozygous) | **11** | 10 | 0 | 0 | 0 | 1 | 0.36 |
|  |  | Autosomal recessive (homozygous) | **4** | 3 | 0 | 0 | 0 | 1 | 1.50 |
|  |  | X-linked | **6** | 6 | 0 | 0 | 0 | 0 | -0.66 |
|  | Inconclusive | Autosomal dominant De novo | **6** | 6 | 0 | 0 | 0 | 0 | -0.66 |
|  |  | Autosomal dominant inherited | **4** | 3 | 0 | 0 | 0 | 1 | 1.50 |
|  |  | Autosomal dominant unknown | **1** | 1 | 0 | 0 | 0 | 0 | -0.27 |
|  |  | Autosomal recessive (compound heterozygous) | **3** | 3 | 0 | 0 | 0 | 0 | -0.47 |
|  |  | Autosomal recessive (homozygous) | **3** | 2 | 0 | 1 | 0 | 0 | 0.74 |
|  |  | X-linked | **2** | 2 | 0 | 0 | 0 | 0 | -0.38 |
|  | Negative | Negative | **237** | 220 | 1 | 2 | 0 | 14 |  |

**Supplementary Table 3:** **Recurrent mutations found among pediatric and prenatal participants.** All the participants with recurrent mutations were unrelated. Kinship analysis using exome sequencing data was conducted using PC-Relate.

| **Gene (Number of recurrent mutation)** | **Nucleotide change and position** | **Inheritance pattern** | **Prenatal/ Pediatric** | **R/E categories reported by parents of participant 1** | **R/E categories reported by parents of participant 2** | **Outcome defined by CSER** |
| --- | --- | --- | --- | --- | --- | --- |
|  |  |  |  | **with recurrent mutations** | **with recurrent mutations** |  |
| *PTPN11* (2) | c.182A>G | Autosomal dominant (De novo) | Pediatric, prenatal. | LT | Missing | Positive |
|  | (Pathogenic) |  |  |  |  |  |
|  | (p.Asp61Gly) |  |  |  |  |  |
| *CDK13* (2) | c.2525A>G | Autosomal dominant (De novo) | Both pediatric | LT | EU, LT, NAT | Positive |
|  | (Pathogenic) |  |  |  |  |  |
|  | (p.Asn842Ser) |  |  |  |  |  |
| *FGFR3* (2) | c.742C>T | Autosomal dominant (De novo), Autosomal dominant unknown (father unavailable) | Both prenatal | EU | EA, EU, Missing | Positive |
|  | (Pathogenic) |  |  |  |  |  |
|  | (p.Arg248Cys) |  |  |  |  |  |
| *PTPN11* (2) | c.854T>C | Autosomal dominant (De novo) | Both prenatal | Missing | EU | Positive |
|  | (Pathogenic) |  |  |  |  |  |
|  | (p.Phe285Ser) |  |  |  |  |  |
