## Supplementary figures and images for "Diagnostic Yield of Exome Sequencing in a Diverse Pediatric and Prenatal Population is not Associated with Genetic Ancestry"

### Supplementary Figure 1

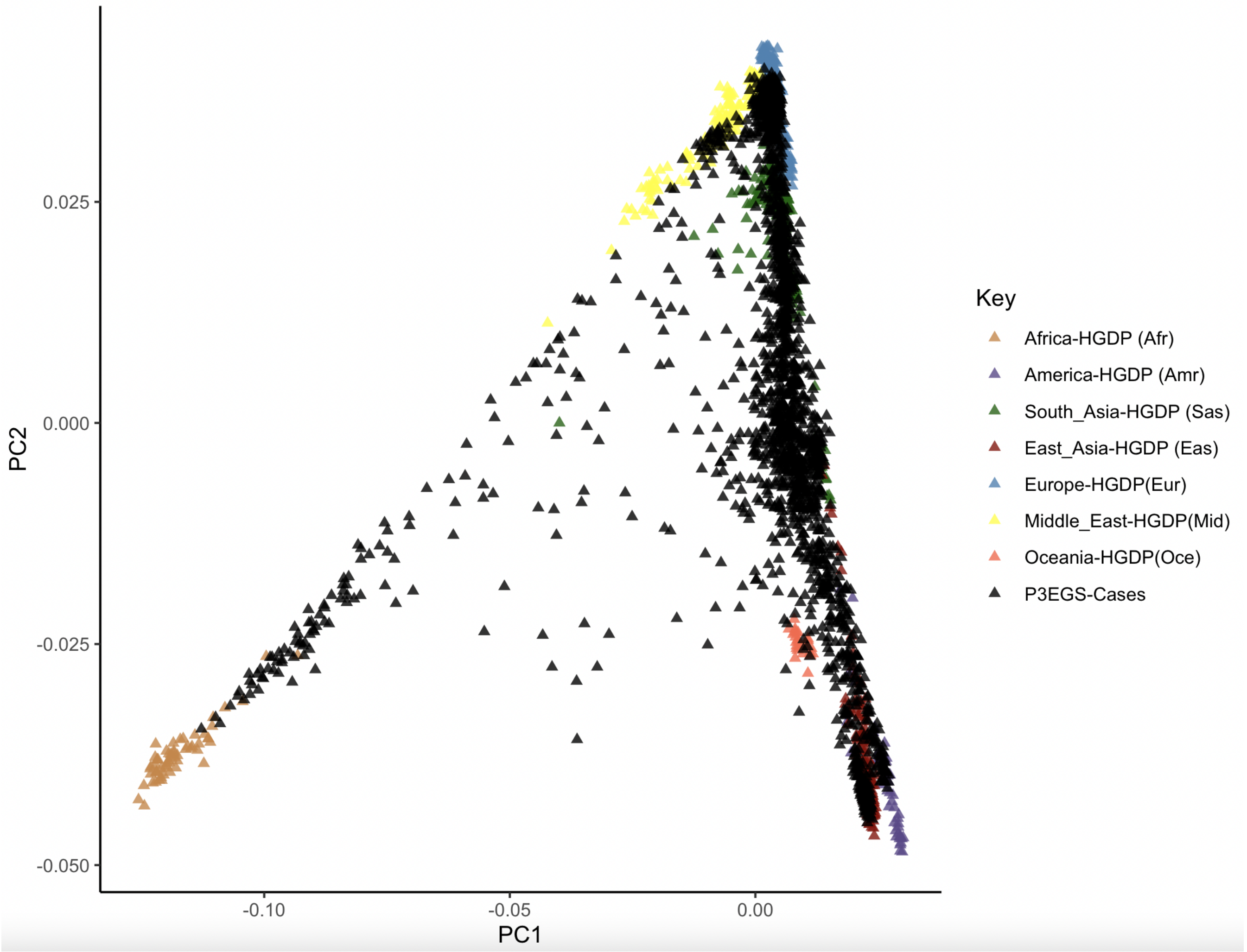

### Supplementary Figure 2

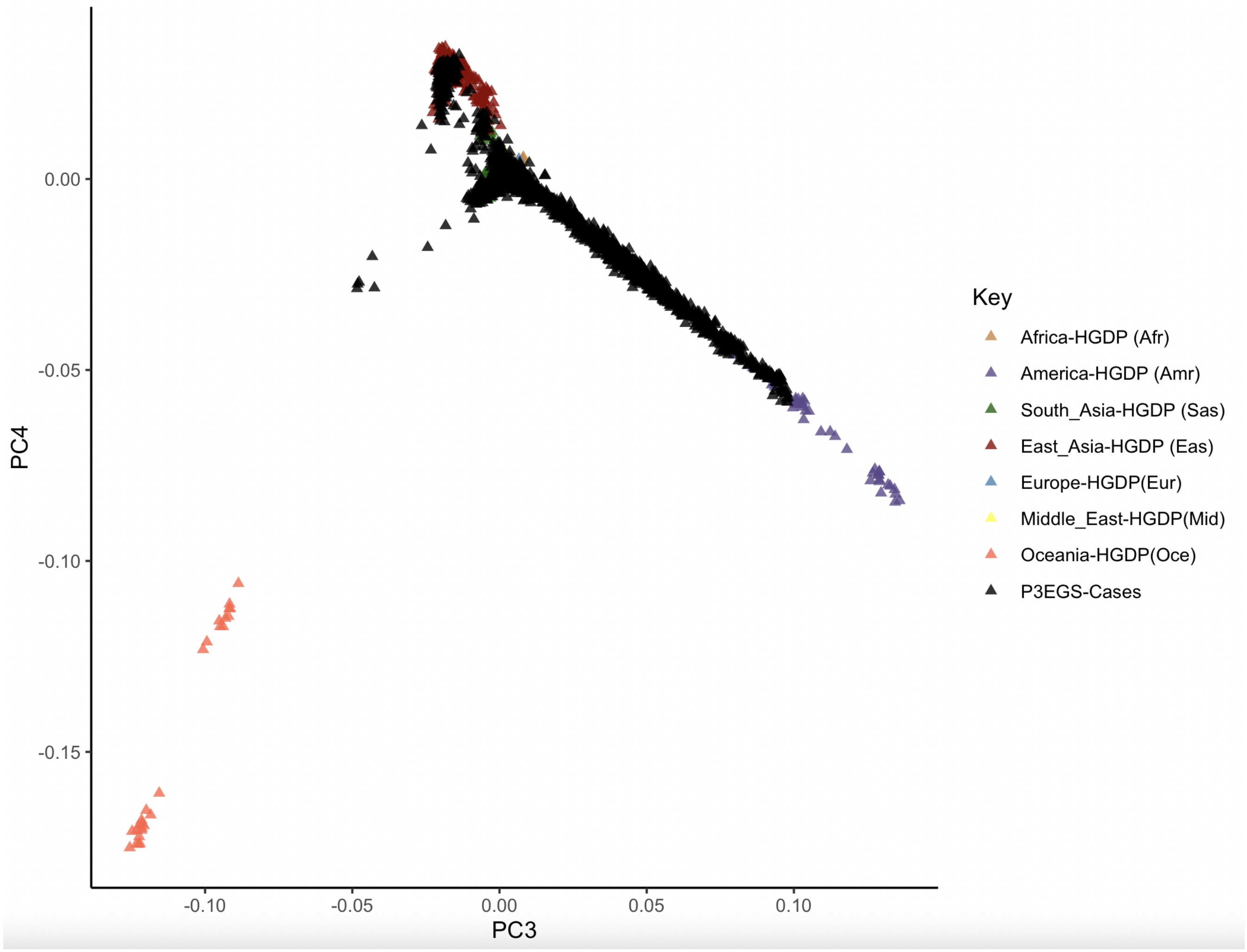

### Supplementary Figure 3

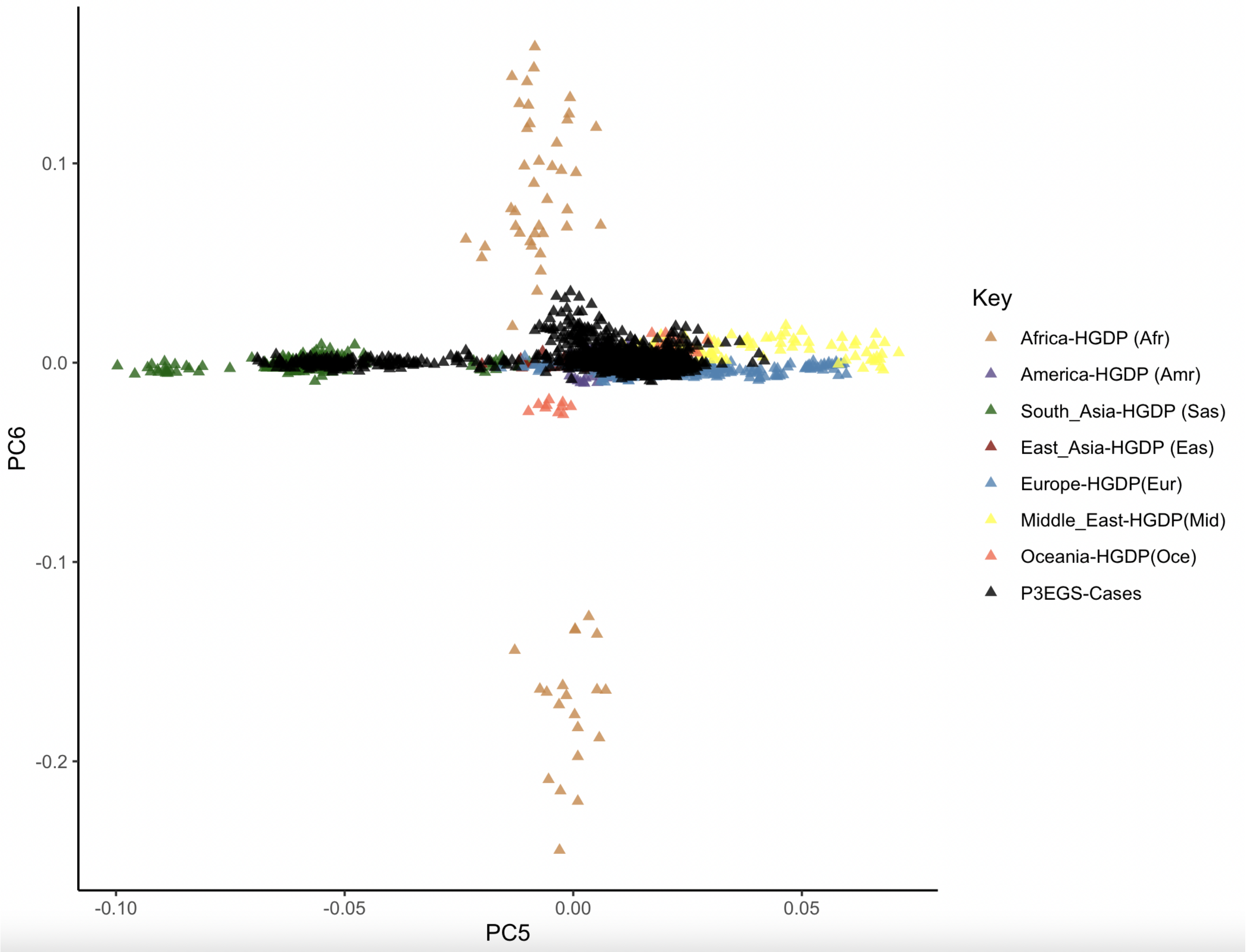
